# Model Based Estimation of the SARS-CoV-2 Immunization Level in Austria and Consequences for Herd Immunity Effects

**DOI:** 10.1101/2021.03.10.21253251

**Authors:** Martin Bicher, Claire Rippinger, Günter Schneckenreither, Nadine Weibrecht, Christoph Urach, Melanie Zechmeister, Dominik Brunmeir, Wolfgang Huf, Niki Popper

## Abstract

Several systemic factors indicate, that worldwide herd immunity against COVID-19 will probably not be achieved in 2021. Vaccination programs are limited by availability of doses, the number of people already infected is still too low to have a disease preventing impact and new emerging variants of the virus seem to partially neglect developed antibodies from previous infections. Nevertheless, after one year of COVID-19 observing high numbers of reported cases in most European countries, we might expect that the immunization level should have an impact on the spread of SARS-CoV-2. We used an agent-based simulation model to reproduce the COVID-19 pandemic in Austria to estimate the immunization level of the population as of February 2021. We ran several simulations of an uncontrolled epidemic wave with varying initial immunization scenarios to assess the effect on the effective reproduction number. We also used a classic differential equation SIR-model to cross-validate the simulation model. As of February 2021, 14.7% of the Austrian population has been affected by a SARS-CoV-2 infection which causes a 9% reduction of the effective reproduction number and a 24.7% reduction of the prevalence peak compared to a fully susceptible population. This estimation is now recomputed on a regular basis to publish model based analysis of immunization level in Austria also including the fast growing effects of vaccination programs. This provides substantial information for decision makers to evaluate the necessity of NPI-measures based on the estimated impact of natural and vaccinated immunization.

## 1 Introduction

According to the WHO chief scientist Dr. Soumya Swaminathan (interview on 2021.01.11, Deutsche Welle TV), herd immunity with respect to the ongoing COVID-19 pandemic will likely not be reached in 2021. Despite increasing availability and roll out of vaccines and (partial) immunization of individuals/persons with a recent infection, the spread of the pandemic cannot be mitigated without further non-pharmaceutical interventions (NPIs) such as social distancing and increased hygienic awareness. Nevertheless, one can expect that an increasing number of people immunized by having been exposed to the virus will at least slightly slow down the current spreading of SARS-CoV-2. We furthermore want to denote this by the term *herd effect*.

To assess this effect, first, the total number of persons already exposed to the virus needs to be estimated. This number generally consists of the cumulative number of detected SARS-CoV-2 infections and the corresponding amount of cumulative undetected infections that occurred in the background. Since the ratio between these cohorts depends on the reporting system, it is very difficult to transfer findings between different countries. The ratio is most often estimated using randomized studies screening for antibodies. But these studies are very time and cost intensive and therefore rarely performed on a regular basis. Other approaches rely on indirect methods such as estimates for the infection fatality ratio and the case fatality ratio [1].

Given the number of persons already exposed to the virus, we evaluate the herd effect using modeling and simulation scenarios. Hereby we will compare the disease spread on the example of fictional epidemic waves with and without immunity. We assume that exposed persons developed sufficient antibodies to protect from secondary infections, but also elaborate the limitations and consequences of this assumption. As outcome measures, we investigate the reduction of the effective reproduction number and of the peak prevalence number.

For both tasks, we apply an agent-based model (ABM) in combination with data from performed cross-sectional SARS-CoV-2 prevalence and serological tests in Austria. This ABM has already been used for several scientific studies such as the investigation of tracing measures [2] or the general dynamics of undetected cases [3], and is one of three models that participate in the official Austrian COVID forecasting panel by the ministry of health (COVID Prognose Konsortium, [4]), which publishes short time forecasts on a weekly basis. For the herd effect, we will also perform cross model validation of the ABM with analytical results from a classic differential equation based SIR-model.

Using this experimental setup, we aim to gain a well founded picture of the total number of known and unknown SARS-CoV-2 cases in Austria until February 2021. We provide estimates on how the corresponding immunization level of the population reduces the maximum epidemiological growth. Based thereon, we formulate the necessary efforts for disease containment.

## 2 Methods

In this section, we present a short introduction to the agent-based model, a specification of the used data and a detailed description of the study setup.

### 2.1 Agent-based simulation model

We use an agent-based model to simulate the COVID-19 pandemic in Austria. In this model, each agent acts as a statistical representative of a member of the Austrian population, characterized by their age, sex, and place of residence. Using a contact network based on households, workplaces, school and leisure time, the disease can be spread from agent to agent. Once infected, an agent passes through different stages of a SARS-CoV-2 infection, including latency period and incubation period. The model also includes a detection probability which defines whether an agent’s infection will later be confirmed by a positive test result and be recorded in the official vigilance system or whether they remain an undetected case. This parameter is a joint summary of different probabilities such as the chance for an asymptomatic disease progression, the chance to be found in a screening program, and the overall test sensitivity used for detection. By simulating each infection and not only the cases represented in the official vigilance systems, the model can provide a complete picture of the epidemic behavior and the total number of people affected by the virus. A more in-depth description of the simulation model as well as used model parameters are provided in parameter tables in [2].

As one of the key features of the agent-based model, the model structure allows a very detailed observation of all processes taking place in the simulation. Consequently, the user has almost no limitations regarding definition of the outcome measures. For the present study, our main interest lies in the dynamics, that is time series, of SARS-CoV-2 infected agents – active, cumulative as well as daily new infected. Since we also investigate the detection process of agents, we also observe the detected agents – active, cumulative as well as daily new detected agents – which correspond to the reported case numbers in Austria. Note, that new detected agents correspond to all agents that were detected (received a positive test) on that specific date. Thus, *new detected* differs from *new infected* not only by a factor (detection probability) but also by a time delay.

### 2.2 Differential Equation SIR model

For cross model validation we compare the results from the agent-based model with results from the classic SIR model by Kermack and McKendrick [5]. It is defined by the three coupled differential equations

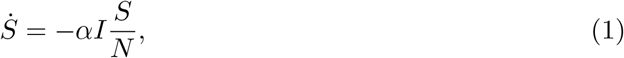

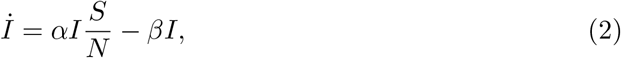

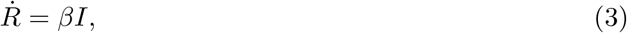

with *N* = *S* + *I* + *R*, and suitable initial conditions. Hereby, the functions *S, I* and *R* stand for the time evolution of the total number of susceptible, infectious, and recovered persons in the system.

### 2.3 Data

In order to parametrize and calibrate the simulation model, a wide range of different data sources has been used. Since these data sources are not specific for the present study, but match the used parameters in previous works, we refer to the parameter tables found in the supplemental material of [2]. With respect to the present study, we want to focus on data sources that reveal information about the detection rate of cases. In Austria, several randomized studies have been performed to estimate the total number of current or past SARS-CoV-2 infections.

In April, SORA Institute for Social Research and Consulting determined that 0.33% (95%CI: 0.12%*−* 0.75%) of the population or approximately 28500 (95%CI: 10200 – 67400) people were infected with SARS-CoV-2 between April 4^th^ 2020 and April 6^th^ 2020 (sample size = 1544) [6]. Statistics Austria performed two screening programs between April 22nd 2020 and April 25^th^ 2020 and between May 26^th^ 2020 and May 30^th^ 2020 [7]. In the first screening program, 1 of 1432 samples was tested positive resulting in a maximum of 0.15% of the population being currently infected (upper bound of the 95%-confidence interval) and in the second screening program, none of the 1279 samples were tested positive.

During the second wave of the COVID-19 pandemic in Austria, Statistics Austria performed once more a randomized screening study [7] and estimated that 3.1% (95%CI: 2.6% *−* 3.5%) of the people aged 16 or older were currently infected with SARS-CoV-2 between November 12^th^ 2020 and November 14^th^ 2020 and that 4.7% (95%CI: 3.8%*−*5.6%) showed antibodies, meaning they had already been infected with SARS-CoV-2 before mid / end of October 2020 (sample size = 2263).

### 2.4 Estimating the immunization level in February 2021

For the estimation of the immunization level, we ran our simulation model for the time span between February 2020 and February 2021. We identified the parameter for the detection rate based on study data and the method published in [3].

In specific, applying the method introduced in [3], the results of a prevalence study (SORA [6]) conducted in April 2020 result in a detection probability of 13% (CI 95%: [2%,86%]) [3]. The second study (Statistics Austria [7]), performed during the second wave of COVID-19, indicates an increase of the detection probability to 35% (CI 95%: [24%, 58%]). In our simulation model, we assumed these values to be characteristic for the corresponding epidemic wave. We set the detection probability parameter to a probability of 13% from February 2020 until August 15^th^ 2020 and gradually increased it to 35% until September 1^st^ 2020. This date was chosen because by mid August, Austria implemented a broader testing regime, starting with the free of charge testing of returnees from vacation destinations classified as risk regions and later introduction of excessive nationwide antigen testing for surveillance [8].

To guarantee that the detected cases in the model match the officially reported case numbers, we proceeded analogous to [2] and [3] and calibrated the infection probability parameter as well as all parameters corresponding to governmental policies for this time period using an iterative bisection method. Hereby, the match of the cumulative detected cases and the officially reported numbers for the positive tests is the calibration target.

Ultimately, in order to obtain an estimate of the immunization level in February 2021, we investigate the final state of our simulation runs. The ratio between the total active and past infected agents and the total number of agents in the model (equivalent to the size of the Austrian population) poses the immunization level and hereby the first major outcome of the model. Note that agents who died (1) from COVID-19, (2) from other causes while being infected, and (3) from other causes after being infected are properly removed by the simulation dynamics and do not contribute to the immunization level.

### 2.5 Estimating the herd effect

It is our aim to assess how the presumably achieved seroprevalence could affect the future progression of the epidemic. To that end, we investigate a fictional, unconfined outbreak of the disease and compare simulation results with different *initial* immunization levels. That means, we assume no implementation of new NPIs, no change in the contact behavior or any other effects on epidemic progression besides decrease of the susceptible population. We will test for sensitivity on the initial immunization (*x*% of the total population) in both models and provide a discussion of the obtained differences and similarities in the used indicators. Moreover, we observe target indicators such as the time-dependent effective reproduction number and the prevalence of infected and detected cases and study them in comparison with a scenario without initially immunized persons.

The effective reproduction number ℛ_eff_(*t*) is understood as the average number of infections that are generated by an individual that gets infected at time *t*. Since this number cannot be measured directly in reality, statistical models for inferring the reproduction factor from available data (e.g. reported incidence) must be used. These methods must take into account disease specific parameters, such as the incubation or the infectious period, implying a certain simplified model of disease progression. The effective reproductive number is suitable to differentiate between epidemic progression (ℛ_eff_(*t*) > 1) and a stable or declining regime (ℛ_eff_(*t*) ≤ 1). In this study, we rely on a statistical approach that is available in the software packages `EpiEstim` [9]. For the configuration of the underlying statistical model (serial interval mean and deviation), we use parameters provided by the Austrian Agency for Health and Food Safety [10]. The same approach is used by authorities and agencies in Austria to provide official estimates of the current reproduction number.

Based thereon, we define the *characteristic reproduction factor* 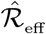 as the reproductive factor 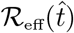 at time 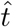 chosen such that (1) the spread of the disease is not yet slowed down by a significant reduction of initially susceptible population and (2) the incidence is high enough to allow for a stable evaluation of the effective reproduction number. Thus, the characteristic reproduction factor is attained in the ‘stable’ phase of an epidemic wave lasting either to the onset of herd immunity effects or until the implementation of countermeasures (i.e. a change in the mixing behavior of the population). Analogously, we assume that a distinct peak/maximum in the observed prevalence is attained at some point in time. We rely on this maximum prevalence number *I*_*max*_ as a second indicator for quantifying the reduction of epidemic spread.

For cross validation, we extract above indicators from two different simulation models that reproduce the fictional wave scenario. The first model is the macroscopic SIR model presented in Section 2.2 and the second model is the ABM introduced in Section 2.1. The former is well suited to generate initial estimates of the investigated target values, since many quantities of the model can be calculated without any numerical simulation. The latter is used to get a more detailed picture of the target measures under consideration of heterogeneous mixing (contact networks).

#### 2.5.1 Herd effect estimation using a classic SIR model

Even though there is no analytic solution to these differential equations, the equations still allow to investigate several important epidemiological measures analytically, as long as no containment related policy is applied that changes the values of *α* and *β* dynamically. For example, ℛ_0_, the basic reproduction rate, is found as the quotient ℛ_0_ = *α/β*. With *S*(*t*) being the population of the susceptible, the effective reproduction rate calculates to

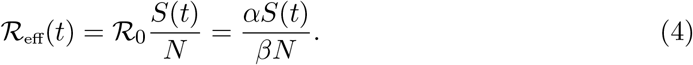

Considering *I* as a function of *S* allows to solve the ODE system and calculate the maximum of the infectious curve by

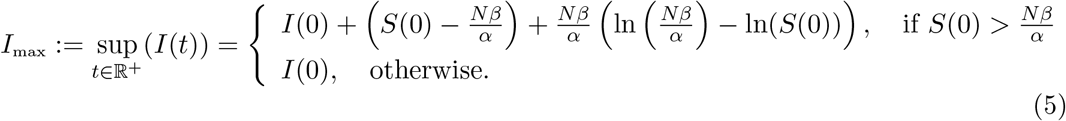

A mathematical derivation of this formula is found in the Appendix.

### 2.5.2 Herd effect estimation using an agent-based approach

We apply the agent-based model to run two sets of scenarios corresponding to the initial household setting of the immune agents. In the first set, we distribute the immune individuals fully randomly in the population. In the second set, we reuse the final state of the simulation used for estimation of the seroprevalence level (see Section 2.4) as the initial state of the new simulation. Hereby, we directly transfer all households with immune agents from the previous simulation and import them into the initial agent population of this simulation. In this way, regional and household clusters from the original simulation are preserved and the spread is more realistic. This approach allows to asses the impact of heterogeneous immunization on our results (compare [11]).

For each scenario, we simulate a hypothetical epidemic wave over the course of six months by randomly introducing 40 infected individuals at the start. This initial number was experimentally determined to produce the most stable results for the stochastic agent-based model.

To calculate the defined outcome measures, we export aggregated daily incidence and prevalence data for the infected and the detected population from our agent-based simulations. The time series of the prevalence allows for direct measurement of the peak height (compare with equation (5)). We further apply the statistical estimator for the effective reproduction number that is provided by the *epiEstim* package on our simulated case data. Hereby, we distinguish between the reproduction calculated from the simulated case incidence 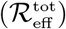 and the one calculated from the detected cases only 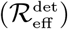. The latter models to the process performed in the real system, the prior is more accurate, but lacks a real world comparator.

## 3 Results

### 3.1 Immunization level

The calibration process of the ABM achieved to match the cumulative detected cases in the simulation with the officially reported case numbers with a root mean square error of *RMSE* = 3721 cases. The quality of the fit can be seen in the lower right part of Figure 1. The other parts of the figure give insights into the dynamics of detected and undetected cases as resulting from the model. Figure 2 shows the share of the population with an active or past infection for selected dates. The selected dates represent the dates relevant for the screening studies (April 5^th^ 2020, mid / end October 2020, November 13^th^ 2020), a date at the end of the first epidemic wave (June 1^st^ 2020) and a current date (February 1^st^ 2021). As of February 1^st^ 2021, 14.1% (97.5%CI: 6.6% *−* 38.1%) of the Austrian population is already immune to SARS-CoV-2 and 14.7% (97.5%CI: 7.1% *−* 38.8%) or approximately 1.3 million people (97.5%CI: 640 000 −3.5 million) has been in contact with SARS-CoV-2 due to an active or past infection. From February 2021 also vaccinated persons will be included in this estimation according to the actual assumptions on immunity and reduction of transmission effects. The corresponding data is available on http://www.dexhelpp.at/en/immunizationlevel. This estimation is recomputed on a regular basis to publish model based analysis of immunization level in Austria also including the fast growing effects of vaccination programs in Austria.

**Figure 1:**
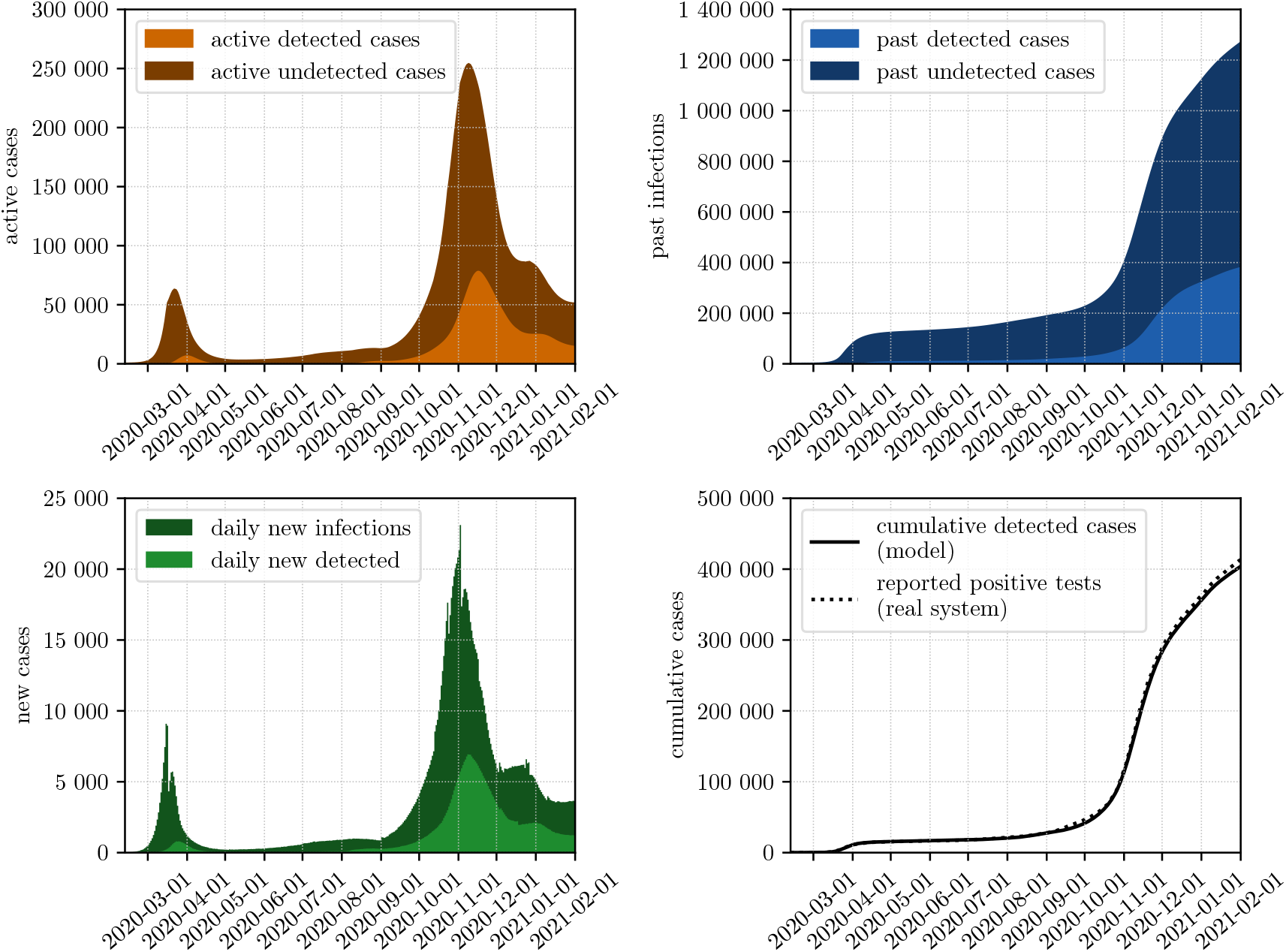
Calibrated model results for the active, past and daily new infections of SARS-CoV-2 from March 12^th^ 2020 until January 12^th^ 2021. Detected infections denote those people who have received a positive test result and are recorded in the official reporting system. The lower right plot displays a comparison between the calibrated model results and the officially reported case numbers.

**Figure 2:**
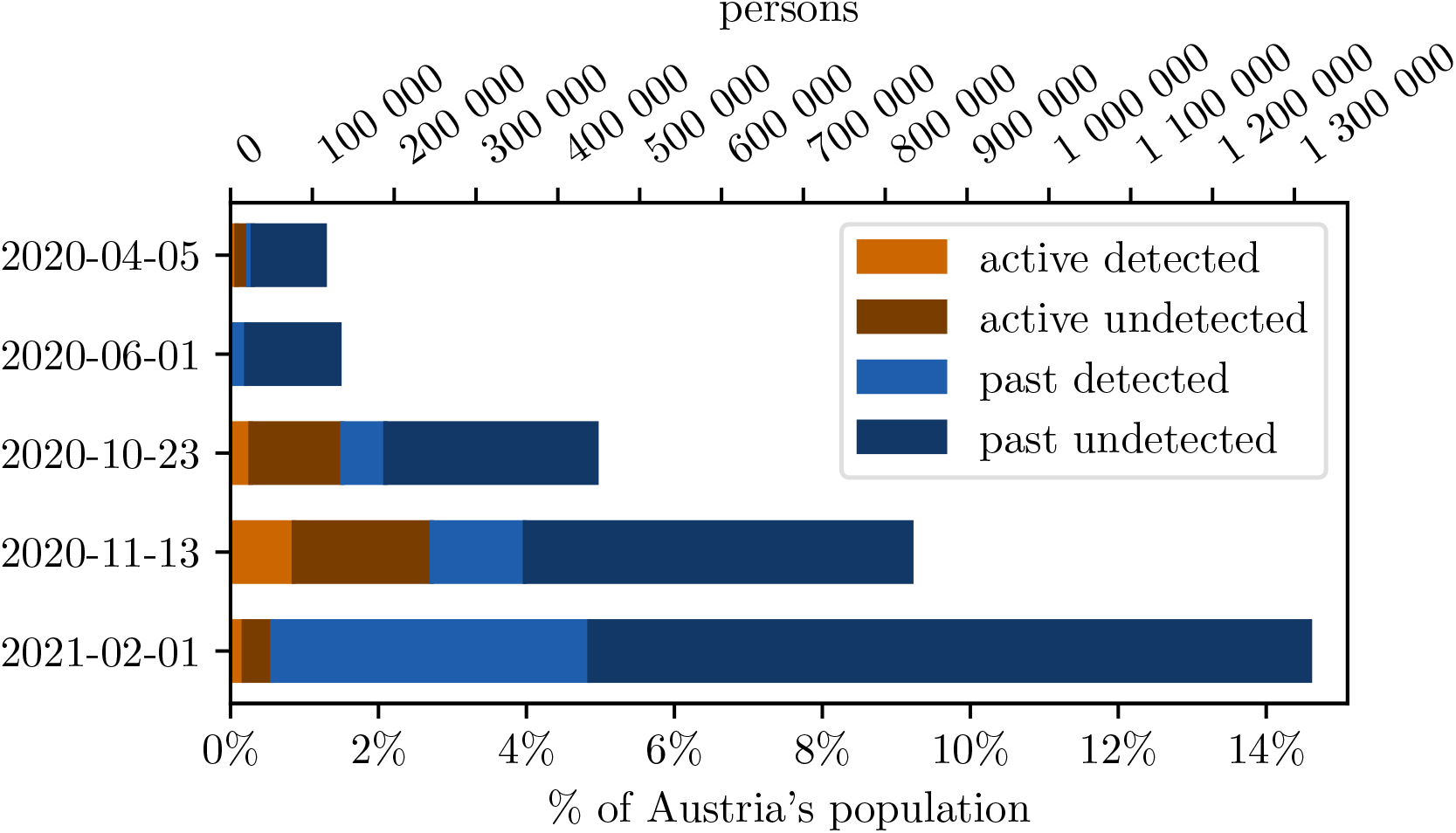
Active and past infections of SARS-CoV-2 for selected dates. Detected infections denote those people who have received a positive test result and are recorded in the official reporting system. The sum of active and past infections denotes the people who have been in contact with SARS-CoV-2 up to that date.

### 3.2 Quantifying the Herd Effect

#### 3.2.1 Classic SIR Model

Assuming that a certain percentage *x* of the population is initially immune (i.e. 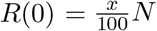), the complementary compartment *S*(0) + *I*(0) = *N − R*(0) is reduced by the same amount. Considering that the initial prevalence *I*(0) is negligible small, 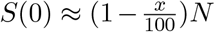 can be derived.

In the classic SIR model, 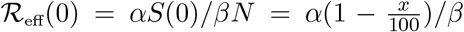. We see that the initial value of ℛ_eff_ depends linearly on the immunity level of the population and 14.1% initial immune persons result in ℛ_eff_(0) being 85.9% of its original value. Note that this statement does not hold for ℛ_eff_(*t*) in general, since the time dynamics of the SIR model are nonlinear.

Moreover, we apply equation (5) to estimate the impact on the peak of the disease wave. For *I*(0) ≈ 0, i.e. (1 *− x*)*S*(0) ≈ (1 *− x*)*N*, we find that

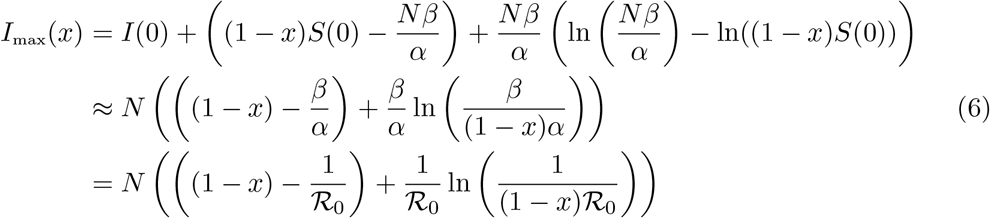

describes the maximum peak height of the epidemic wave, with initially *xN* immune persons. If we investigate the relative reduction of the immunization level 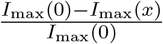, the total population *N* cancels out of the equation. Applying the formula with typical estimate ℛ_0_ = 3, an immunization level *x* of 7.05% results in a reduced peak by 15.4%. Moreover, 14.1% provide a 30.1% reduced peak, 21.15% lead to a 44.0% reduced peak and 28.2% initial immune reduce the maximal peak height by 57.1%. ℛ_0_ = 4 leads to a relative reduction of 13.8% for an immunization level of 7.05%, 25.5% for an immunization level of 14.1%, 37.7% for an immunization level of 21.15% and 49.4% for an immunization level of 28.2%. (see Figure 6). Easily seen, the relation is almost linear for values *x* close to zero.

**Figure 6:**
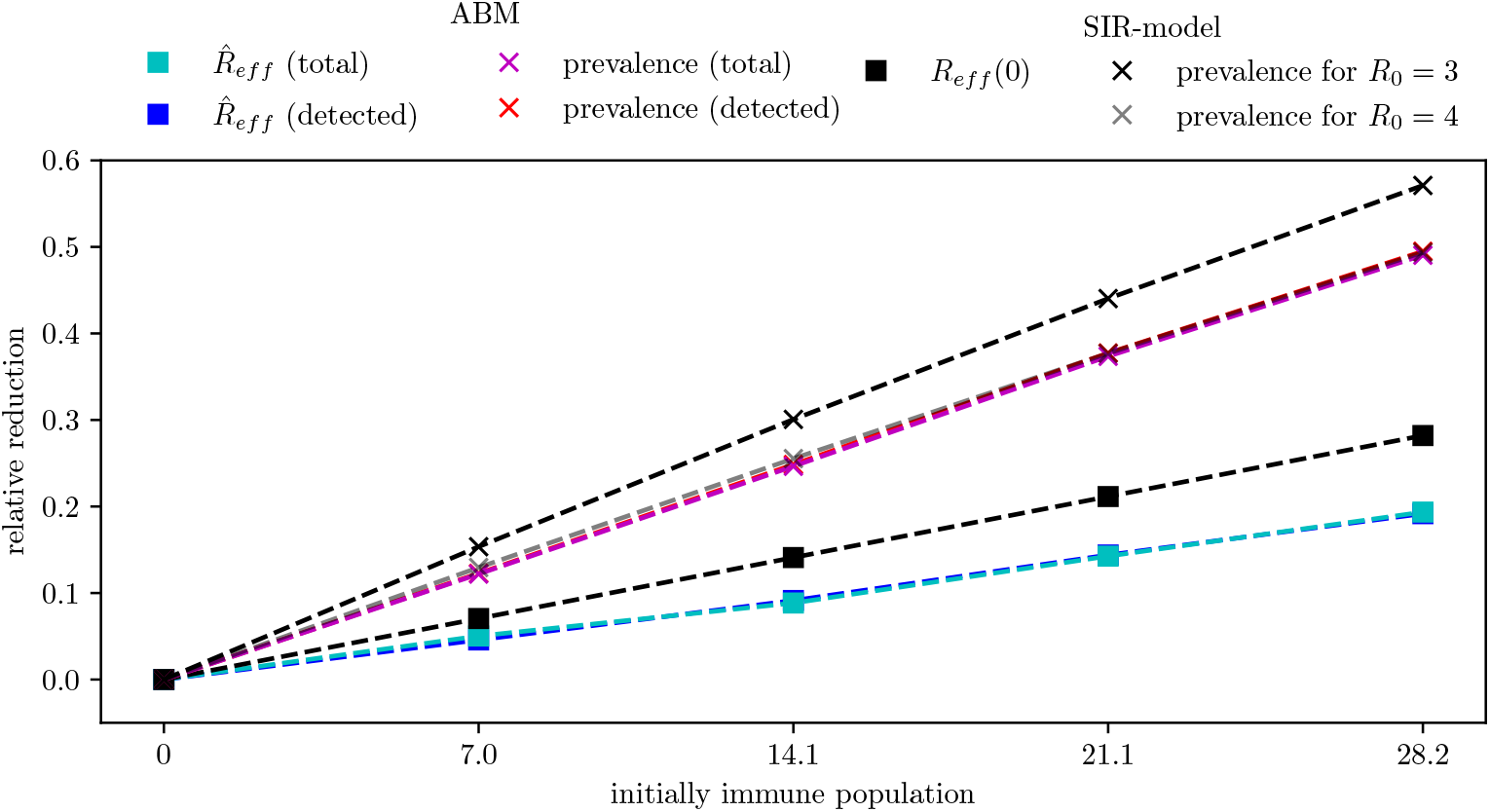
Relative reduction of the effective reproduction number and the peak of the prevalence compared to a fully susceptible population for the results of the classic SIR-model and of the agent-based simulation model

#### 3.2.2 Agent-based model

Using the agent-based model, we simulated 10 different scenarios of an epidemic wave of SARS-CoV-2 infections with different initial network structures and 5 levels of sero-prevalence. Figure 3 depicts various outcome measures of the different scenarios, such as timelines of prevalence and incidence for all cases as well as the detected cases. It can be seen, that a higher immunization level does not only reduce the peak of maximal prevalence but also delays it by 4 days (for 7% immune) to 20-23 days (for 28.2% immune). Furthermore, for all scenarios, the timeline of the new detected cases is delayed by about 9 days compared to the timeline of new infections. This delay occurs because cases are detected about 9 days after their infection as the mean incubation time is 5.1 days and the mean reaction time between symptom onset and positive test results is 3.8 days [3]. The detected cases are also reduced to 35% of all cases which corresponds to the detection probability applied for these scenarios. The network structure of the initially immune individuals has only a small effect on the epidemic curve. Incidences are delayed by 1-2 days with the delay growing with the immunization level but the overall dynamic and the height of the incidence peak is not affected otherwise. Therefore, in the following, we will only present the results for the scenarios with randomly distributed initially immune individuals. The results for the other scenarios can be found in the appendix.

**Figure 3:**
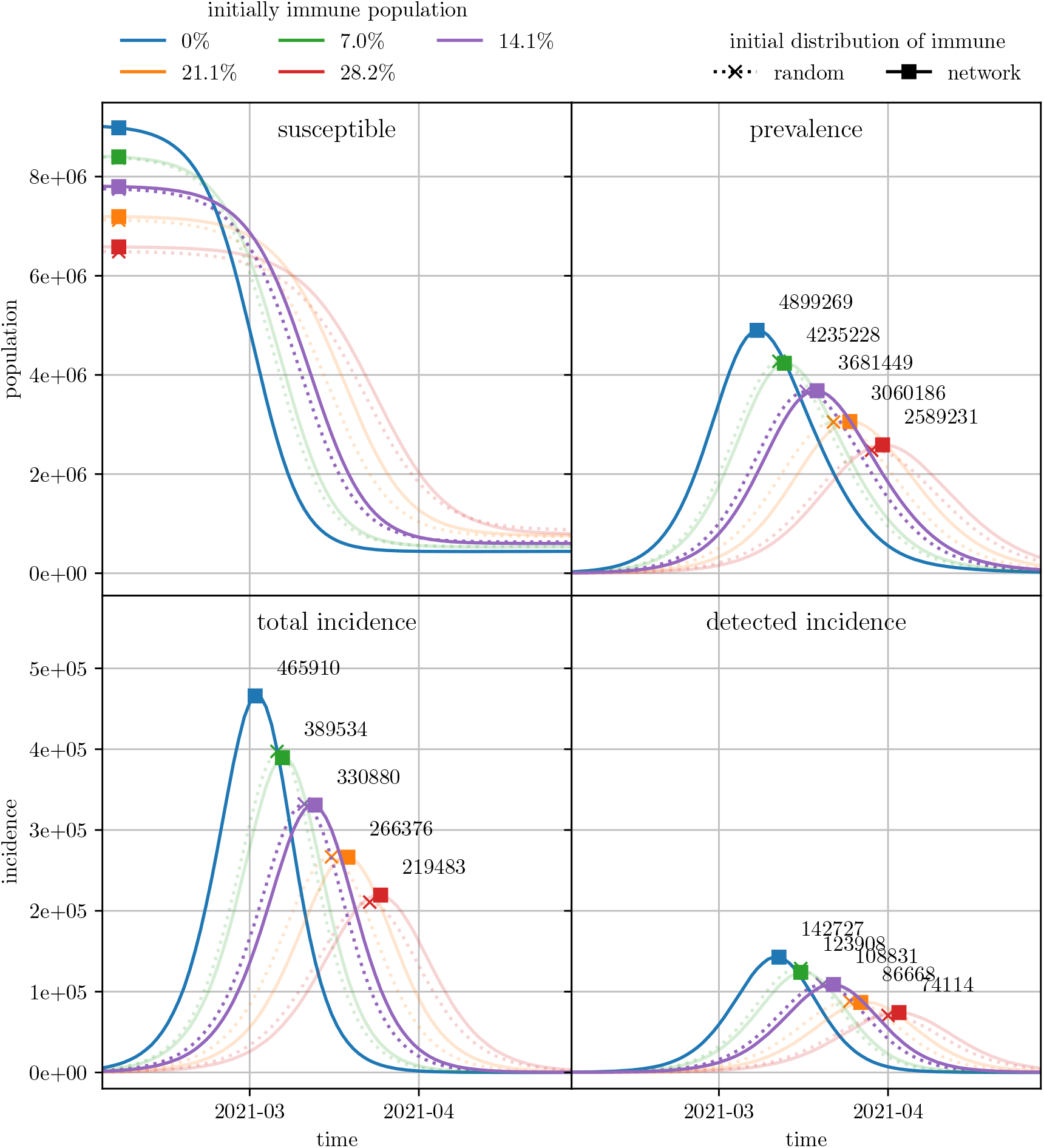
Various outcome measures for scenarios with different initially immunization levels. For all scenarios with varying immunization level and distribution of initially immune individuals, the number of susceptible (upper left) and infected (upper right) individuals is depicted. Daily new incidences are evaluated for all cases (lower left) and detected cases (lower right). For each timeline, the maximum of the curve is marked.

Figure 4 depicts the temporal dynamic of the effective reproduction number in the different scenarios. In the lower part, 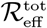 has already been shifted by 9 days to account for the delay between infection and detection date. After this temporal shift, 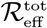 and 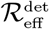 can be considered as equivalent [9]. Especially in the early phase of the epidemic wave, where the disease can spread uninhibited, the two measures correspond very well and both variants of ℛ_eff_ reach the threshold value of 1 at the same point in time.

**Figure 4:**
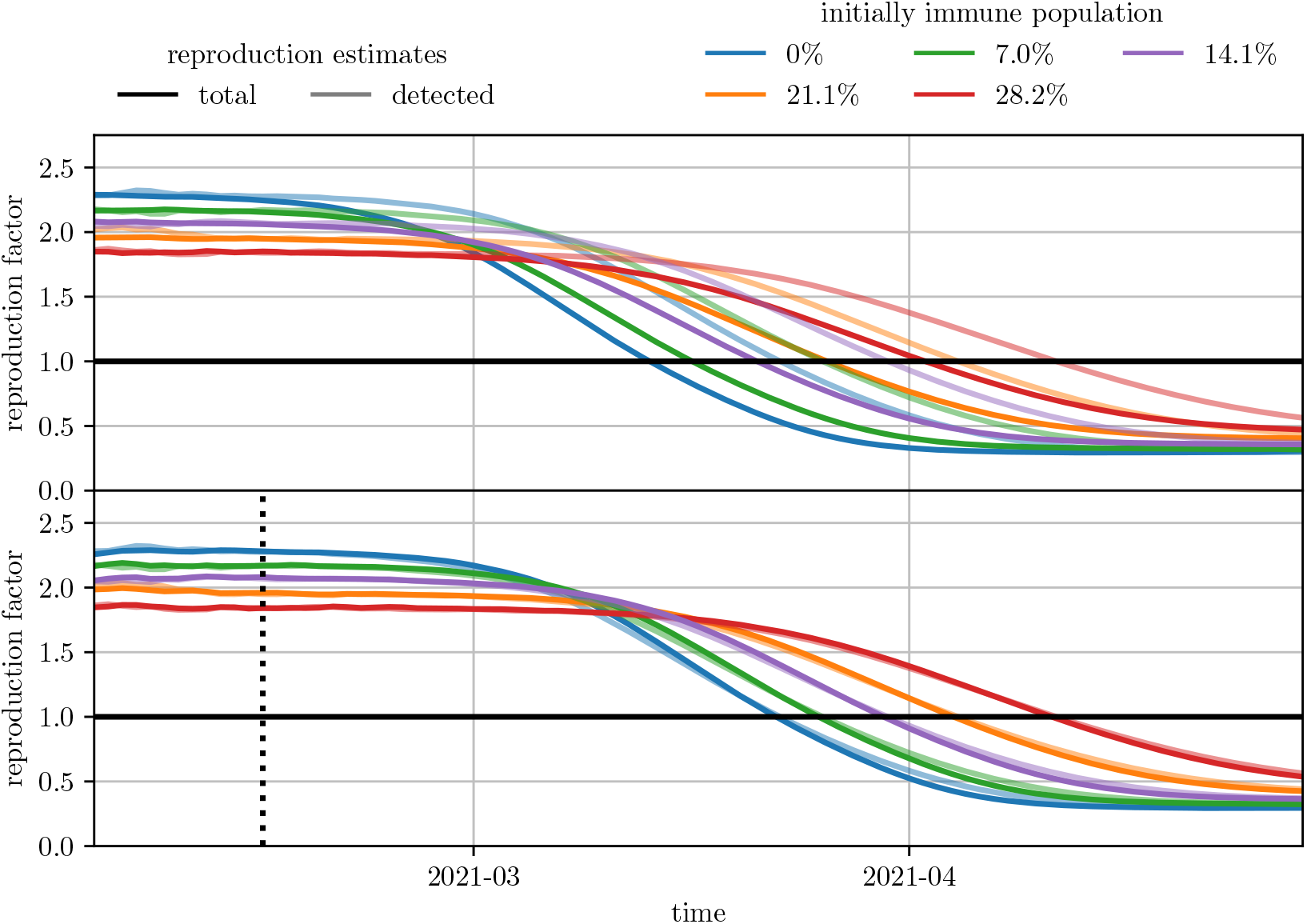
Estimates of the effective reproduction number for different initial immunization level. The effective reproduction number has been evaluated both on the total new cases as well as on the detected new cases. In the lower plot, the evaluation based on all new cases has been shifted to account for the time delay between infection and detection date. The black dotted line indicates the date (2021-02-14) used for evaluation of 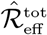 and 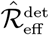.

The values of 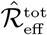 and 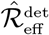, as well as the maximal prevalence, are depicted in Figure 5. Moreover, Figure 6 shows the relative reduction of these outcome measures compared to the scenarios with an initially fully susceptible population. For the results of the agent-based simulation model, the relative reduction of the outcome measures is independent on whether the outcome measures were evaluated based on all infections or only on the detected cases. The reduction of 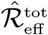 and 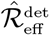 caused by a higher level of initial immunization follows the linear trend as in the classic SIR-model. However, in the agent-based model, the relative reduction caused by an initial immunization level of 14.1% lies only between 8.8% (for detected cases) and 9.1% (for all cases). The relative reduction of the maximal prevalence matches well with the estimates generated with the classic SIR-model and ℛ_0_ = 4.

**Figure 5:**
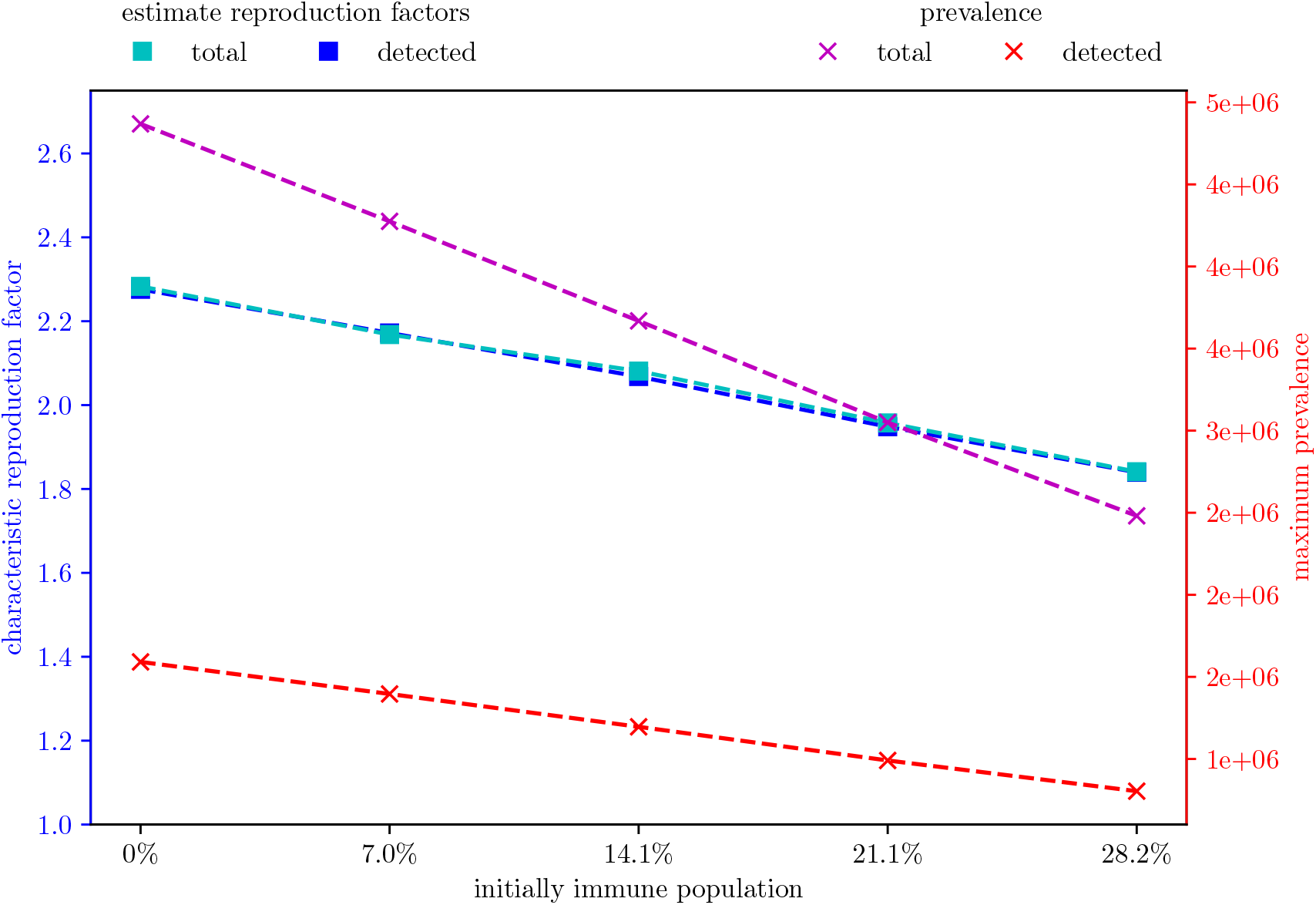
Effective reproduction number 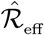 and maximal prevalence as a function of initial immunization level for the results of the agent-based simulation

## 4 Discussion

An agent-based model was used to give a detailed estimate on the level of immunization among the Austrian population in February 2021 and how this influences the future spread of the pandemic. The modeling and simulation study reveals that with February 1^st^ 2021, a total number of 1.328 million Austrian inhabitants have already been exposed to the virus of which 1.319 million are still alive to this date, 7778 died due to COVID-19, and 1030 died after having recovered from the disease. Assuming that the persons being exposed to the virus developed sufficient antibodies for immunization, 14.1% of the Austrian population can be considered immune by February 2021. The results of the modeling study indicate that this level of immunization leads to a maximum reduction of ℛ_eff_ by 9% compared to a fully susceptible population and a reduction of the maximum prevalence by 24.7% (see Figure 6). Assuming that the virus remains as infectious as it has been in spring 2020, this percentage directly maps to the policies required for containment of the disease.

With respect to quantifying the herd effect, we defined different outcome measures giving similar, yet slightly different pictures. This underlines, that defining consistent outcome measures for assessment of epidemic spread related measures or effects is still a challenging task. In specific, our study shows that even though estimations of the relative reduction of the effective reproduction number due to an initial immunization level can be drawn from the classical SIR-model, these estimations do not directly relate to the results of more complex simulation models. First, the classical SIR-model neglects or oversimplifies some important factors of the modeling of SARS-CoV-2, the most important being the incubation and latency period, the isolation of symptomatic cases and the general stochasticity of the system. Second, measuring ℛ_eff_ in a post-processing step from case numbers gives different results than directly measuring ℛ_eff_ from the model parameters since additional parameter assumptions need to be made.

Apart from these differences, comparisons between the results of the agent-based model and the analytic calculations from the classical SIR model show interesting similarities and insights. First of all, the agent-based model results follow the linearity predicted by the analytic calculations: the maximum value of ℛ_eff_(*t*), approximated by the characteristic reproduction rate, and the maximum peak height of the active cases *I*_*max*_ relate indirect proportionally to the base immunity level of the population. This feature contributes to the cross-model validation of the agent-based model. Secondly, the agent-based model also provides a much deeper insight into the disease spread related processes than the classic SIR model. For example, we find different sources for time-delays in different outcomes such as the delay of about 9 days between 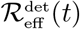 and 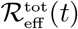 (see Figure 4) and the delay of about 1-2 days caused by a different initial network structure (see Figure 3). The prior nicely depicts why any kind of COVID-19 countermeasure can only show effects in the reported case numbers at the earliest one week after introduction of the policy and is well known among policy makers. The latter is a less obvious observation and requires deeper analysis. It shows that the disease requires about 1 to 2 days to “break out” of naturally grown immunized networks while it can already spread with higher velocity if the initial network of immunized agents is randomly created. This can be emphasised as a surprising finding of this study, since we would have expected a higher impact of the initial network structure.

Our method to evaluate the total number of people who have been in contact with SARS-CoV-2 differs from other approaches using estimates of the infection fatality rate [1] and resulting in an estimated immunization level of 7% for Austria in February 2021. Moreover, our method to quantify the effect of this immunization level is vastly different to methods applied for other infectious diseases that rely on observational studies performed before and after vaccination of a large portion of the population [12] [13].

The results of this study are limited by general limitations of modeling and simulation based research including modeling simplifications and data quality. Like all models on the total amount of infected persons also this work relies on international and national studies and analysis of not detected cases based on measured data. In addition we assumed that a past infection of SARS-COV-2 will always result in immunity long enough for the scope of this study. Studies have shown that antibodies against SARS-COV-2 can be found 6 months after an infection in 84% of the cases [14] and that a previous infection lowers the risk of a new infection by 83% [15]. However, more and more reports of reinfections with SARS-COV-2 emerge with the second wave of COVID-19 since fall 2020, indicating that this model assumption might be too general. Yet, since there is still too little data available, we found the evidence not convincing enough to change fundamental parts of the model at the moment. Furthermore, we did not account for the rising number of mutations of SARS-COV-2. As with general reinfections, we did not consider subsequent infections with different mutations. Similarly, we did not vary the overall infectivity of SARS-COV-2. This means, we did not account for potentially higher infectivity of new mutants that slowly replace current strains.

The data sources used for this study also have several limitations. Some of the randomized prevalence studies have been performed at a time with low case numbers (end of April 2020 and May 2020). Here, the sample size of 1432 and 1279, respectively, were not big enough to produce a significant outcome useful for further analyses. Moreover, all prevalence studies only included people age 16 or older and did therefore provide no further insight in the number of undetected cases among children. Finally, the official data used for calibrating the model comes with a great number of reporting bias. Reported cases vary highly with weekends and national holidays or the testing regime applied in the different Austrian federal states.

Next steps include constant reevaluation of the immunization level and its impact on the spread of COVID-19 in Austria, especially focused on the effect of the vaccination program in Austria. This provides substantial information for decision makers to evaluate the necessity of NPI-measures based on the estimated impact of natural and vaccinated immunization at a given moment. Further research will include additional analysis and adaption of modelling parameters once new evidence from randomized screening studies and more reliable data on the risk of reinfection with new virus strains or the role of waning antibodies is available.

## Data Availability

The data for the simulated SARS-CoV-2 seroprevalence level in Austria are publicly available and are updated regularly (see Link 1).
Data for estimation of the seroprevalence level were drawn from the official Austrian epidemiological reporting system for SARS-CoV-2 (EMS, see Link 2) and the results from (serological and prevalence) randomized screening studies performed in Austria (see Links 3 and 4).
Metadata used for parametrization of the simulation model is found the parameter-tables found in the supplemental of publicly available preprint "Evaluation of Contact-Tracing Policies Against the Spread of SARS-CoV-2 in Austria - An Agent-Based Simulation" (Link 5).

http://www.dexhelpp.at/en/immunization_level/

https://covid19-dashboard.ages.at/

https://www.sora.at/nc/news-presse/news/news-einzelansicht/news/covid-19-praevalenz-1006.html

https://www.statistik.at/web_de/statistiken/menschen_und_gesellschaft/gesundheit/covid19/index.html

https://www.medrxiv.org/content/10.1101/2020.05.12.20098970v4

## Acknowledgements

This work was supported by the Austrian Federal Ministry for Social Affairs, Health, Care and Consumer Protection.

## Funding information

This work was funded by the Austrian Research Promotion Agency (FFG) COVID-19 Emergency Call, the Vienna Science and Technology Fund WWTF-COVID-19 Rapid Response Funding, the Medical Scientific Fund of the Mayor of the City of Vienna and the Society for Medical Decision Making (SMDM) COVID-19 Decision Modeling Initiative.

## A Derivation of Formula (5)

We consider the classic SIR model (1)-(3) with ℛ_0_ = *α/β* > 1, initial conditions *I*(0), *S*(0), *R*(0) > 0, and *I*(0) + *S*(0) + *R*(0) = *N*.

For the first case, we will investigate *S*(0) > *Nβ/α*. This ensures, that *I* ^*′*^ (0) > 0 and the results of the model will show a classic epidemic outbreak.

Since equation (1) has a strictly negative right hand side, the resulting function *S* must be monotonically decreasing. Consequently, *S*^*−*1^ is a well defined function on the value set of *S* and we may investigate *F* defined by

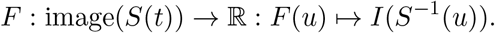

With *u* = *S*(*t*) the function results in *F* (*S*^*−*1^(*S*(*t*))) = *I*(*t*) and therefore describes the number of the infectious *I* as a function of the susceptible *S*.

Using the chain rule, and the rule for the differentiation of inverse functions, we derive

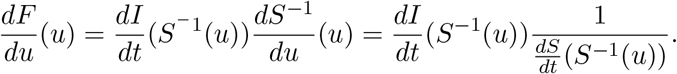

Applying the classic SIR equations (1-3) we follow

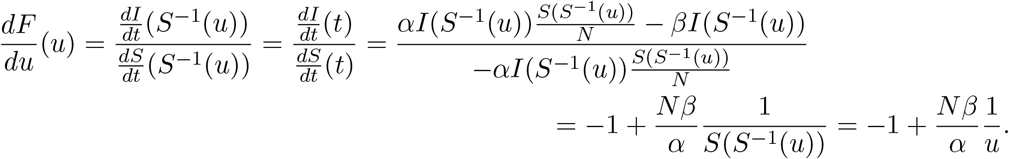

Since

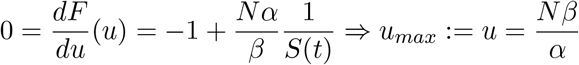

and

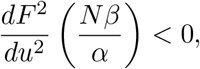

the value 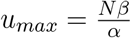 maximizes *F*, as long as it lies in the image of *S*(*t*). This is necessarily true, since *S*(0) > *u*_*max*_ and *I* ^*′*^ (*t*) > 0 for all *S*(*t*) ′ *u*_*max*_. Therefore, *S* needs to drop below *u*_*max*_ in the course of the epidemic wave and there exists a *t*_*max*_ > 0 s.t.

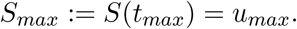

Finally, we may find the corresponding maximum peak value *I*_max_ = *I*(*t*_*max*_) = *F* (*S*(*t*_*max*_)) via integration:

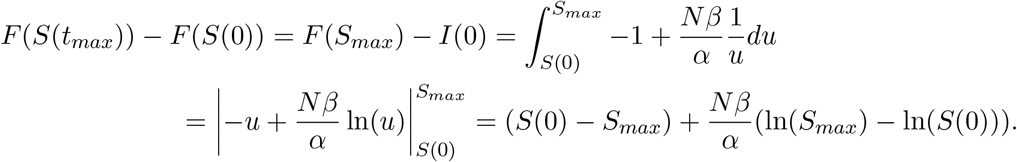

Therefore

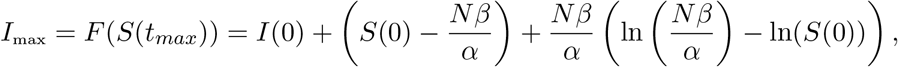

for the first case with 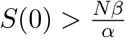.

For the second case with 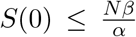, we find *I*^*t*^(*t*) *<* 0 for all *t* > 0 leading to a monotonically decreasing function *I*(*t*). Consequently, *I* has its maximum at *t* = 0.

## B Results for Scenario with Network

Figures 7 to 9 display the analogous curves as shown in Figures 4 to 6 for the scenarios with a model-grown initial immunization network.

**Figure 7:**
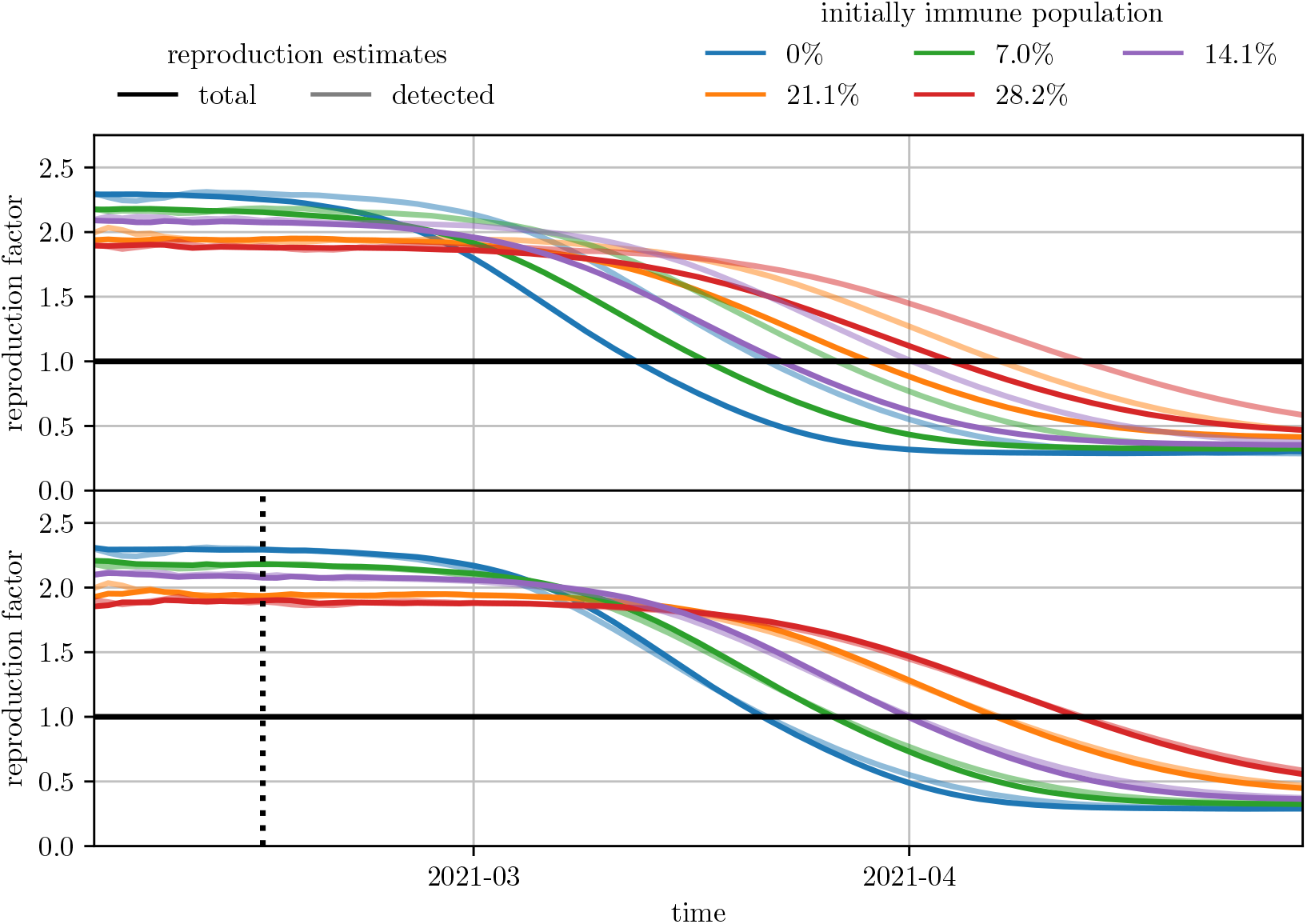
Estimates of the effective reproduction number for different initial immunization level. The effective reproduction number has been evaluated both on the total new cases as well as on the detected new cases. In the lower plot, the evaluation based on all new cases has been shifted to account for the time delay between infection and detection date. The black dotted line indicates the date (2021-02-14) used for evaluation of 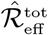 and 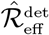.

**Figure 8:**
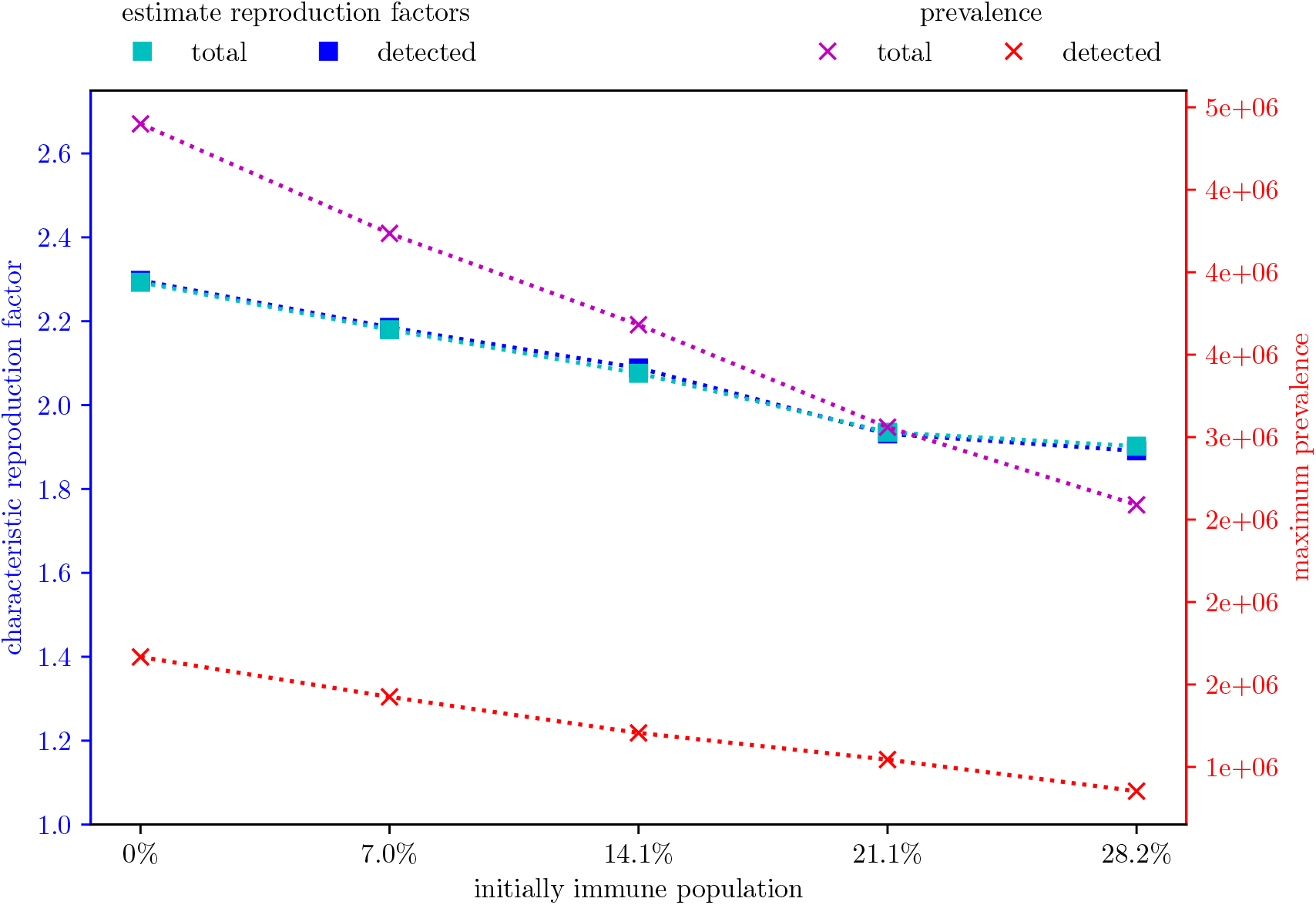
Effective reproduction number and maximal prevalence as a function of initial immunization level for the results of the agent-based simulation

**Figure 9:**
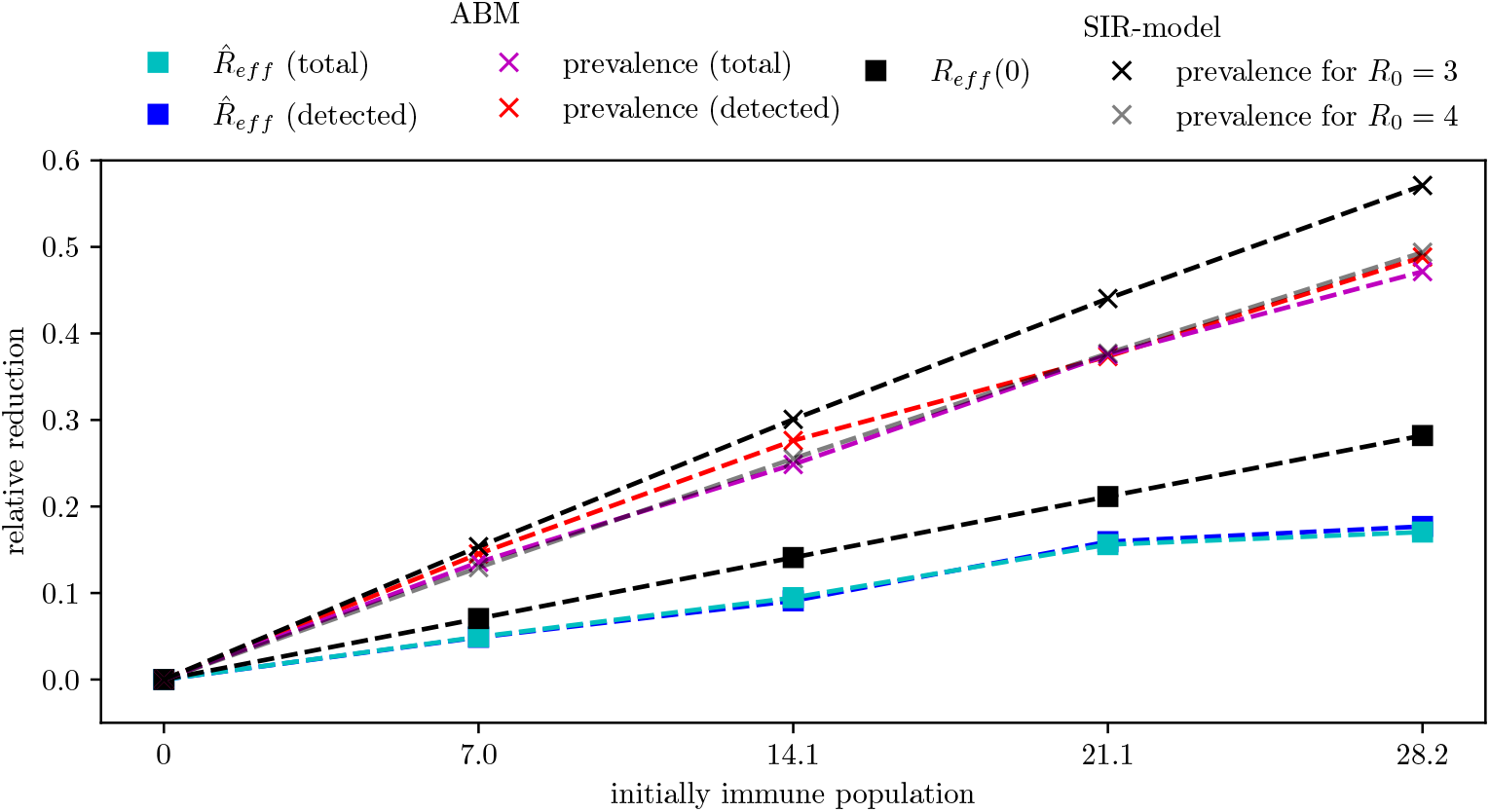
Relative reduction of the effective reproduction number and the peak of the prevalence compared to a fully susceptible population for the results of the classic SIR-model and of the agent-based simulation model

## Notes

### Competing Interest Statement

The authors have declared no competing interest.

### Author Declarations

The study had no need of any ethics committee approval

## References

[1] M. Sáanchez-Romero, V. di Lego, A. Prskawetz, and B. L. Queiroz, “An indirect method to monitor the fraction of people ever infected with covid-19: An application to the united states,” PloS one, vol. 16, no. 1, p. e0245845, 2021.

[2] M. R. Bicher, C. Rippinger, C. Urach, D. Brunmeir, U. Siebert, and N. Popper, “Agent-based simulation for evaluation of contact-tracing policies against the spread of SARS-CoV-2,” p. 2020.05.12.20098970, publisher: Cold Spring Harbor Laboratory Press. [Online]. Available: https://www.medrxiv.org/content/10.1101/2020.05.12.20098970v2

[3] C. Rippinger, M. Bicher, C. Urach, D. Brunmeir, N. Weibrecht, G. Zauner, G. Sroczynski, B. Jahn, N. Mühlberger, U. Siebert, and N. Popper, “Evaluation of undetected cases during the COVID-19 epidemic in austria,” vol. 21, no. 1, p. 70. [Online]. Available: https://doi.org/10.1186/s12879-020-05737-6

[4] M. Bicher, M. Zuba, L. Rainer, F. Bachner, C. Rippinger, H. Ostermann, N. Popper, S. Thurner, and P. Klimek, “Supporting austria through the covid-19 epidemics with a forecast-based early warning system,” medRxiv, 2020. [Online]. Available: https://www.medrxiv.org/content/early/2020/10/20/2020.10.18.20214767.1

[5] W. O. Kermack and A. G. McKendrick, “A contribution to the mathematical theory of epidemics,” Proceedings of the royal society of london. Series A, Containing papers of a mathematical and physical character, vol. 115, no. 772, pp. 700–721, 1927.

[6] SORA institut: Corona-virus dunkelziffer. [Online]. Available: https://www.sora.at/nc/news-presse/news/news-einzelansicht/news/corona-virus-dunkelziffer-1027.html

[7] COVID-19 prävalenz. [Online]. Available: https://www.statistik.at/webde/statistiken/menschenundgesellschaft/gesundheit/covid19/index.html

[8] Bundesministerium für Soziales, Gesundheit, Pflege und Konsumentenschutz, “Antigen-tests im rahmen der Österreichischen teststrategie sars-cov-2,” Bundesministerium für Soziales, Gesundheit, Pflege und Konsumentenschutz, Tech.

[9] Rep., Oct 2020. [Online]. Available: https://www.gesundheit.gv.at/r/krankheiten/immunsystem/coronavirus-covid-19/FactsheetAntigen21.10.2020.pdf

[10] A. Cori, N. M. Ferguson, C. Fraser, and S. Cauchemez, “A new framework and software to estimate time-varying reproduction numbers during epidemics,” American journal of epidemiology, vol. 178, no. 9, pp. 1505–1512, 2013.

[11] L. Richter, D. Schmid, A. Chakeri, S. Maritschnik, S. Pfeiffer, and E. Stadlober, “Schätzung des seriellen intervalles von COVID19, Österreich,” p. 3.

[12] G. Schneckenreither and N. Popper, “Dynamic multiplex social network models on multiple time scales for simulating contact formation and patterns in epidemic spread,” in 2017 Winter Simulation Conference (WSC). IEEE, 2017, pp. 4324–4335.

[13] S. L. Pollard, T. Malpica-Llanos, I. K. Friberg, C. Fischer-Walker, S. Ashraf, and N. Walker, “Estimating the herd immunity effect of rotavirus vaccine,” Vaccine, vol. 33, no. 32, pp. 3795–3800, 2015.

[14] M. Drolet, É. Bénard, M.-C. Boily, H. Ali, L. Baandrup, H. Bauer, S. Beddows, J. Brisson, J. M. Brotherton, T. Cummings et al., “Population-level impact and herd effects following human papillomavirus vaccination programmes: a systematic review and meta-analysis,” The Lancet infectious diseases, vol. 15, no. 5, pp. 565–580, 2015.

[15] D. Ladage, D. Röosgen, C. Schreiner, D. Ladage, C. Adler, O. Harzer, and R. J. Braun, “Persisting antibody response to sars-cov-2 in a local austrian population,” medRxiv, 2020. [Online]. Available: https://www.medrxiv.org/content/early/2020/11/23/2020.11.20.20232140

[16] V. Hall, S. Foulkes, A. Charlett, A. Atti, E. Monk, R. Simmons, E. Wellington, M. Cole, A. Saei, B. Oguti, K. Munro, S. Wallace, P. Kirwan, M. Shrotri, A. Vusirikala, S. Rokadiya, M. Kall, M. Zambon, M. Ramsay, T. Brooks, C. Brown, M. Chand, and S. Hopkins, “Do antibody positive healthcare workers have lower sars-cov-2 infection rates than antibody negative healthcare workers? large multi-centre prospective cohort study (the siren study), england: June to november 2020,” medRxiv, 2021. [Online]. Available: https://www.medrxiv.org/content/early/2021/01/15/2021.01.13.21249642

